# Patient insights research exploring disease awareness, patient life experience, and current management of visceral leishmaniasis in Bihar, India

**DOI:** 10.1101/2024.07.11.24309649

**Authors:** Shyam Sundar, Fabiana Alves, Koert Ritmeijer, Margriet den Boer, Colin Forsyth, Bethan Hughes, Clare Zamble, Kirsten Carter, Gerhild Angyalosi

## Abstract

**Background:** Visceral leishmaniasis (VL) is a vector-borne disease caused by *Leishmania* parasites and transmitted by sand fly bites, targeted for elimination in India. VL primarily affects rural, low-income populations with limited health care access. In South Asia, few studies have explored patients’ perspectives, diagnoses, and treatment experiences, and how these experiences influence their expectations about treatment.

**Methodology/principal findings:** A qualitative study was conducted in Bihar, India, using moderator-facilitated, protocol-defined discussion. Eighteen adult patients and 12 caregivers of children diagnosed with and treated for VL within the last 12 months were identified by self-report. Mean time from symptom onset to diagnosis was 13.8 days. Challenges of the patient life experience included lack of urgency by health care professionals, delayed diagnosis, and unavailability of current standard-of-care therapy where a patient receives their diagnosis (63% switched to a different center for treatment, at times delaying treatment). Key barriers include out-of-pocket financial burden, absence from work/home duties, and long-distance travel to hospitals. Patients expressed a preference (29/30) for a potential oral treatment that could be taken close to home over the current standard of care (infusions/injections in clinics).

**Conclusions/significance:** This study reveals new insights and confirms previous research showing access to care for patients with VL in the Bihar area remains a challenge. Although most patients with VL seek care early, diagnosis often requires multiple visits to a health care facility. Despite access to therapy in public hospitals, a substantial number of patients prefer private clinics. Even in public locations, many patients need to move from the diagnostic center to another center to receive therapy, creating an additional burden for patients. As a potential alternative to current parenteral treatment, oral therapy would be preferred by adult patients and caregivers of pediatric patients and may reduce barriers to access to care.

**AUTHOR SUMMARY:** This is an interview-based investigation of patient awareness, experience, and individual challenges for adults and caregivers of children seeking diagnosis and effective care for visceral leishmaniasis (VL) in rural districts of Bihar, India. Patients’ descriptions of their health experiences revealed obstacles at every stage, including limited information about the illness at the community level, financial burdens associated with loss of work, and cost of travel to seek diagnosis. Although our study was subject to recall bias, it elicited insights from the patients’ own experiences and revealed deterrents and burdens that may not have been visible otherwise. Our study also found that most patients would prefer a potential oral treatment for VL over the current parenteral–based standard of care. This patient-driven approach of identifying unmet needs and potential gaps in public health access can be useful in furthering the aims of the National Elimination Program for VL in India and in other areas with endemic VL, particularly those planning to introduce elimination programs.

## INTRODUCTION

Visceral leishmaniasis (VL), also known as kala-azar, is a vector-borne disease found in 90 countries in the tropics, subtropics, and southern Europe, in settings ranging from rain forests to deserts [1]. In India, VL is endemic among 54 districts across 4 states of Bihar, Jharkhand, West Bengal, and Uttar Pradesh, leaving a population of approximately 140 million people at risk of infection [2].

VL was targeted for elimination in India by 2020 [3], and in the last decade, several major policy changes have been implemented with the goal of elimination. Reported cases of VL have been consistently decreasing in India, from 16.82 cases per 100,000 in 1990 to 0.60 cases in 2019 [4, 5]. The most recent provisional government data showed that in 2023 there were 520 VL cases nationwide and only 4 deaths [6], indicating control of the disease and potentially putting the goal of eliminating VL as a public health problem within reach. Provisional government analysis shows it is likely that all 633 subdistricts endemic for VL achieved elimination target (of <1 VL case per 10,000 population at the sub-district level [7]) in 2023 [8]. Elimination of VL is a high priority in India, with government monetary incentives in place to encourage detection and diagnosis in the community, and to assist patients with obtaining treatment [9]; however, little is known about patients’ perspectives, diagnoses, and treatment experiences, and how these experiences influence their expectations about treatment.

In the Indian subcontinent, VL is caused by *Leishmania donovani* parasites; human transmission occurs via the sand fly *Phlebotomous argentipes*, without known animal reservoirs [10]. Symptoms of VL include irregular bouts of fever, weight loss, liver and spleen enlargement, and anemia; untreated, VL is fatal in >95% of cases [3]. Post kala-azar dermal leishmaniasis (PKDL) is a cutaneous disease occurring in areas of India where VL is prevalent and can appear months or years after a patient has been treated for VL or sometimes in patients with no past VL history [5, 11]. Patients with chronic PKDL can be a reservoir for sustained VL transmission, and thus are an important factor in achieving total VL elimination [5].

VL is mostly prevalent in rural areas and affects primarily low-income populations with limited healthcare access [10]. Prevalence is associated with malnutrition, population displacement, poor housing, a weak immune system, and poverty [3]. In a previous study in Bihar, residence in highly marginalized communities was found to be the greatest risk factor for being affected by VL [12].

In India, the current standard-of-care therapy for VL consists of 10 mg/kg single-dose liposomal amphotericin B by intravenous infusion [13, 14]. Although highly efficacious, infusions are challenging in resource-limited settings, requiring drug refrigeration and close monitoring of patient renal function and electrolyte levels [15]. Because access to medication is challenging in the areas where VL is most prevalent, even when tests and medicines are free of charge, roughly three-quarters of households affected by VL in Bangladesh [16, 17] and Bihar [13] experience financial hardship in obtaining diagnosis and treatment due to factors such as travel costs and missed work.

A few studies on knowledge of and attitudes toward VL, barriers to care and reporting, and costs of patient management have been conducted in Bihar, other parts of India, and Nepal [18–22]. One key single-center, cross-sectional study conducted at a research hospital in Bihar evaluated the patient quality of life (QoL) associated with VL in India using the abbreviated quality of life survey from the World Health Organization (WHO BREF) QoL tool [23]. VL was shown to have a significant negative impact on physical, psychological, social relationship, and environmental domains of QoL in 95 patients. In a separate study, semi-structured, in-depth interviews explored QoL among 29 patients with VL treated at a medical institute in Bihar who were coinfected with HIV [24]. This study also reported substantial negative impact on patients, especially economically. Despite these initial findings, deeper insights are needed, particularly into the early part of the patient life experience before patients have been admitted to the kinds of research hospitals in which previous studies have been conducted. These insights could provide a more comprehensive understanding of the VL patient experience.

In response to this unmet need and in the context of the National Elimination Program, we aimed to understand perspectives of people affected by VL who had sought care in their communities by identifying and recruiting study participants from their homes, rather than from a research or treatment facility. The stated objectives of our qualitative study were as follows: to gain insights regarding patients’ experience of the disease, diagnosis, and treatment; to better understand current levels of disease awareness and barriers in diagnosis and treatment access; and to explore the acceptability of an oral treatment formulation among adult patients and caregivers of pediatric patients. The results of our research provide useful insights into challenges and barriers reported by patients seeking treatment in a historical area of high VL prevalence.

## METHODS

### Study Design

A qualitative study was conducted using moderator-facilitated discussion, with a predefined set of screening criteria and following a protocol-defined discussion guide. Institutional Review Board (IRB) ethics approval was received from Sigma-IRB (https://www.sigma-irb.com/) on December 28, 2022. Recruitment took place in districts across Bihar, the region that accounts for the majority of VL cases in India, clustered into 3 groups: 1) Vaishali, Gopalgunj, and Saran; 2) Saharsa, Madherpura, Araria, Purnia, and Katihar; and 3) Muzaffarpur, East Champaran, Sitamarhi, and Samastipur. The data report is shared in **Supporting Information 1.**

Interviewers visited district hospitals to identify villages/townships that were hotspots of patients with VL in the past 24 months (per study eligibility criteria). Recruiters met with local pharmacists, community health care professionals (HCPs), and local leaders/village council members to confirm the relevance of target areas. Recruitment was conducted door-to-door, where consent was gained for screening. Patients were recruited via a screening questionnaire to determine eligibility and willingness to take part in the interview. If written or audio-recorded consent was given, respondents could be screened face-to-face at that time or at a later appointment arranged at the respondent’s convenience. Sixty-minute, face-to-face, in-depth interviews were conducted in Hindi by a local partner agency, which were transcribed, translated, and analyzed in a standardized way using an analysis grid, to identify patterns and frequency of mention of key themes and datapoints.

Inclusion criteria consisted of the following: adult patients aged ≥18 years or parents/legal guardians of pediatric patients aged <18 years, self-reported as having been diagnosed with VL and treated in the last 2 years, and from a mix of agricultural and nonagricultural backgrounds, with ≤50% having agricultural work as their main occupation. All participants recruited had been diagnosed and treated within the last 12 months. The screening process aimed for an equal distribution between genders among adult patients and a mix of ages among pediatric patients: ≤4 children aged <5 years and ≥8 children aged 5 to 17 years. The sample size was chosen to balance feasibility and reasonable robustness for analysis.

Fieldwork began in February/March 2023. After review of a partial sample (22 completed interviews), it was noted that there was little consistency in the types of treatments the patients recalled being given. In addition, some patients reported being treated in the private sector, which was an unexpected finding. Because patients were unable to name the treatments that they received and many appeared to have received multiple treatments during their illness, the researchers felt it necessary to add a disease validation measure into the protocol. Recruitment was paused in June 2023 and the protocol was resubmitted to the IRB and approved June 14, 2023 to include required proof of confirmed diagnosis either as documentation (ie, VL treatment card, blood test result, doctor’s letter) or details of an HCP or Accredited Social Health Activist (ASHA; a community health worker employed by India’s Ministry of Health and Family Welfare), who would be able to confirm VL diagnosis and/or treatment. In July 2023 recruitment recommenced, and previously recruited patients were requested to provide documentation to support their diagnosis and treatment. The 8 patients recruited after the revision of the protocol were all required to provide a VL treatment card.

Data were collected using the discussion guide in Hindi, consisting of 5 sections: context and patient/caregiver activation; patient journey overview; patient journey: initial symptoms to diagnosis; patient journey: treatment and follow-up; and potential new VL treatment (**Table 1**).

**Table 1.**
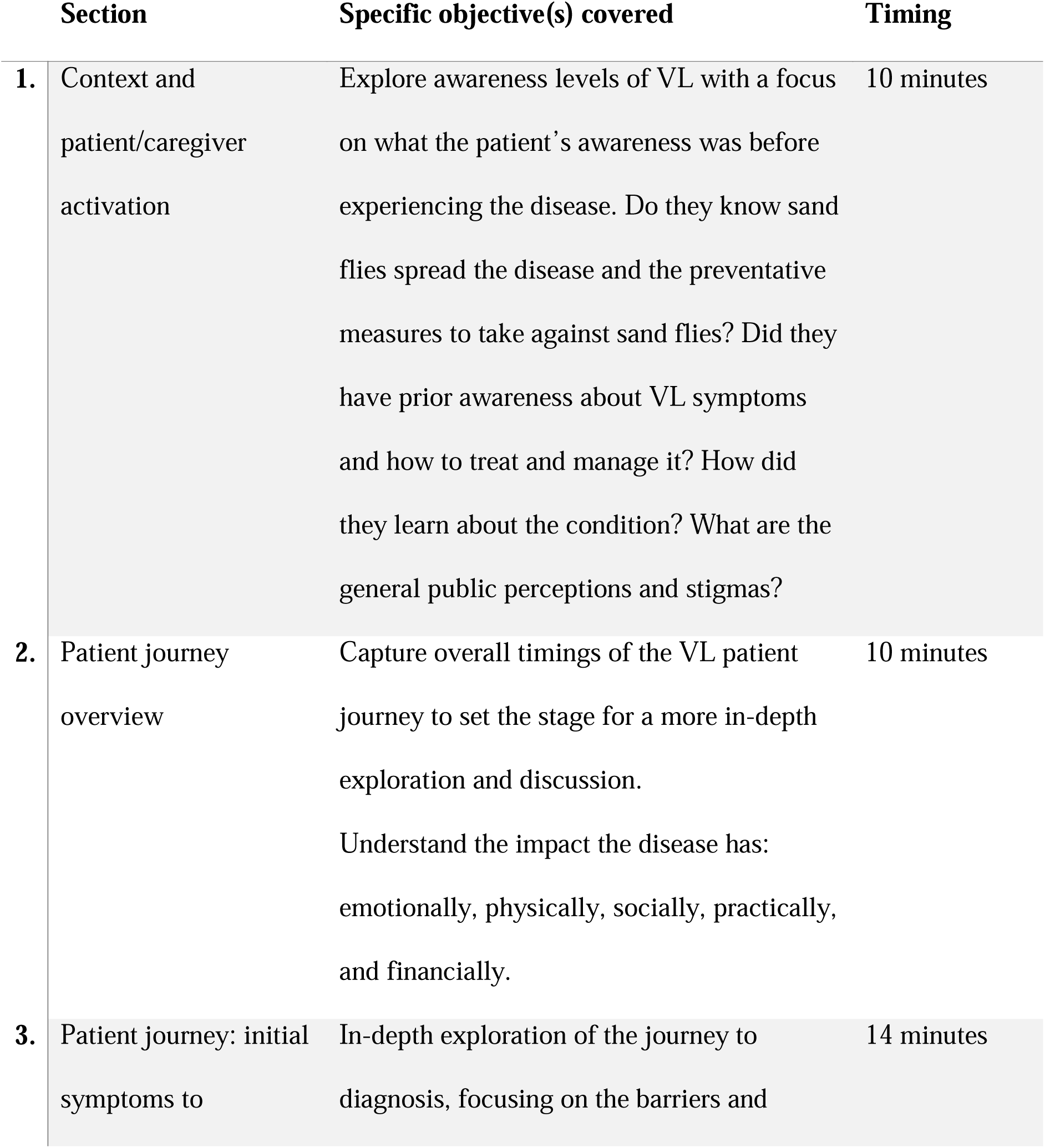
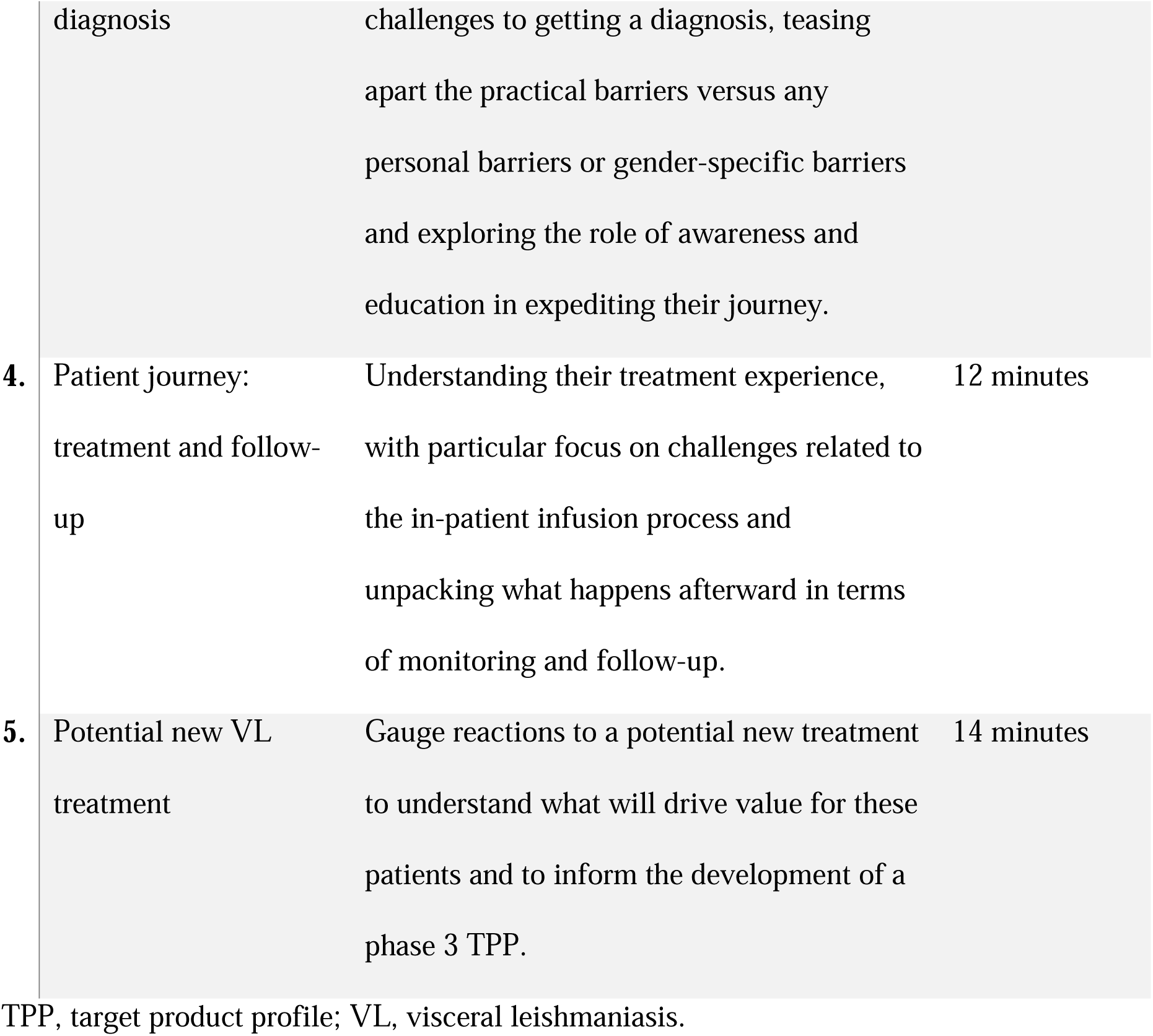
Patient Interview Discussion Guide.

## RESULTS

### Participants

Eighteen adult patients and 12 caregivers of children who self-reported being diagnosed and treated for VL within the last 12 months were included (N=30; **Table 2**). Inclusion criteria allowed for treatment within the last 24 months, but all had been diagnosed and treated within the previous 12-month period. Twelve out of 30 participants or caregivers validated diagnosis and treatment with a VL card; 13 out of 30 had lost or no longer possessed a VL card; and 5 out of 30 were unavailable for recontact for retrospective validation. Patients were located in approximately 25 dispersed villages throughout the region (**Fig 1**), with a maximum of 2 respondents interviewed per village. Most patients were farmers (n=15) and homemakers (n=10), 19 (63%) had received a secondary/higher secondary school certificate or above, and 23 (77%) reported income representing working class and above categories (**Table 2**). This group includes 9 patients who were from the middle lower class, with an annual income of $6,000 to $18,000 (□500,000-□1,500,000) for the entire family.

**Fig 1.**
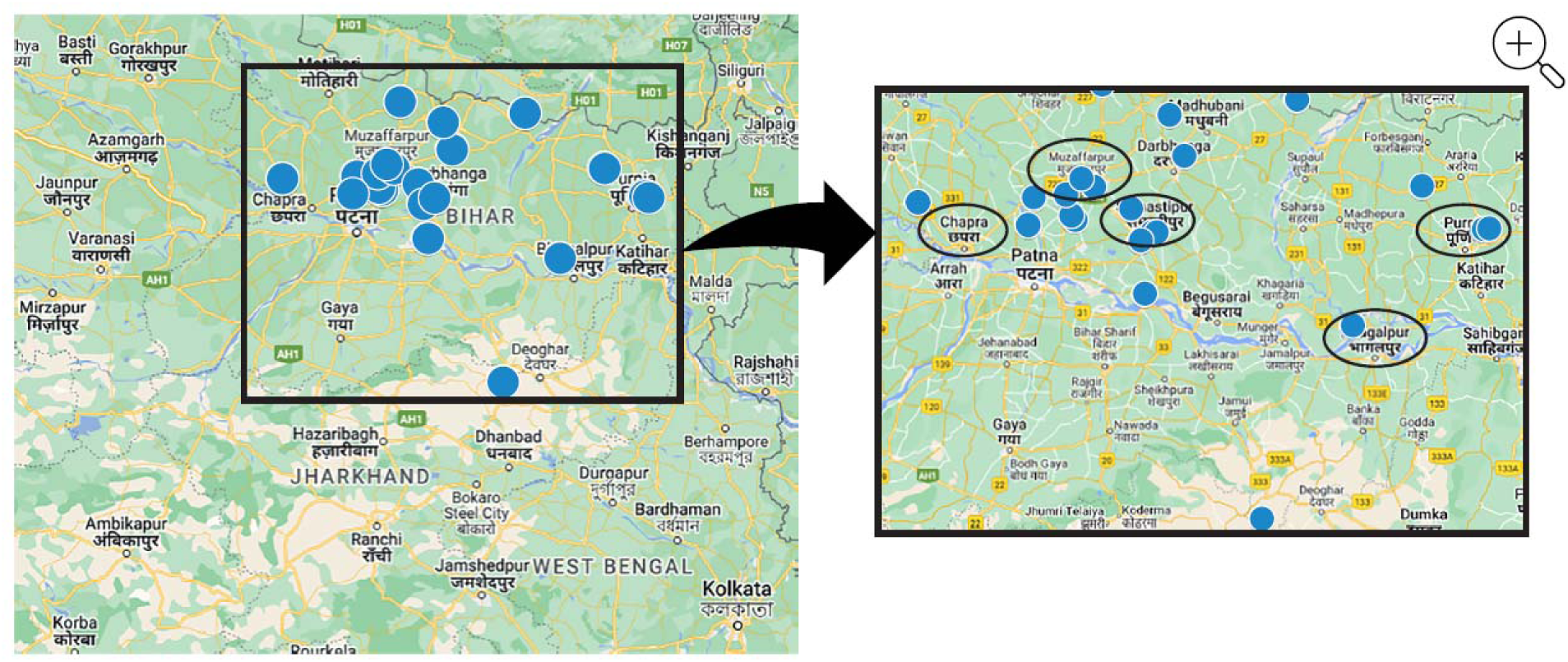
Geographic locations of interviews.^a^. ^a^Numbers denote the approximate location of respondent villages, not individual respondent identification numbers. Maximum 2 respondents interviewed per location.

**Table 2.**
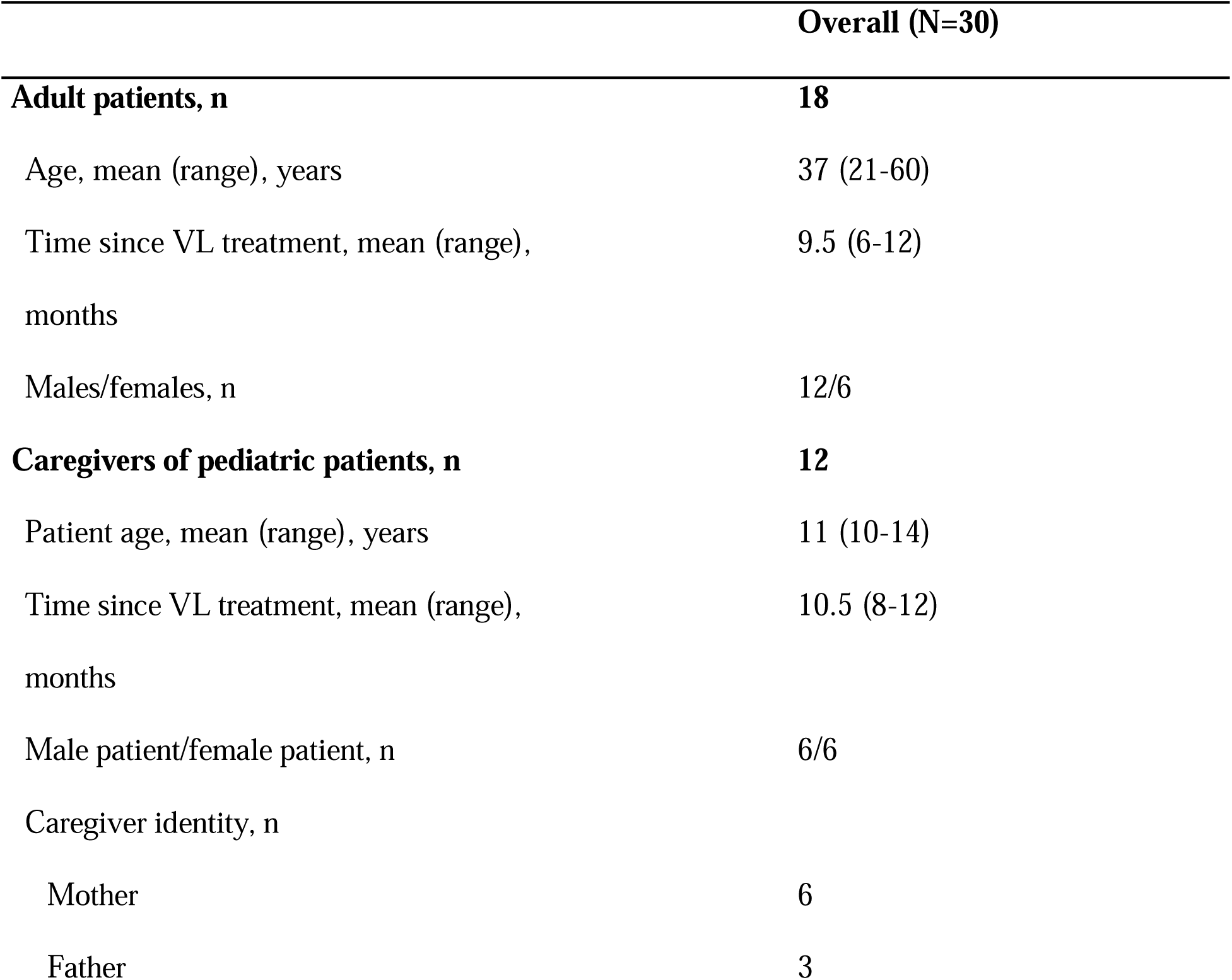
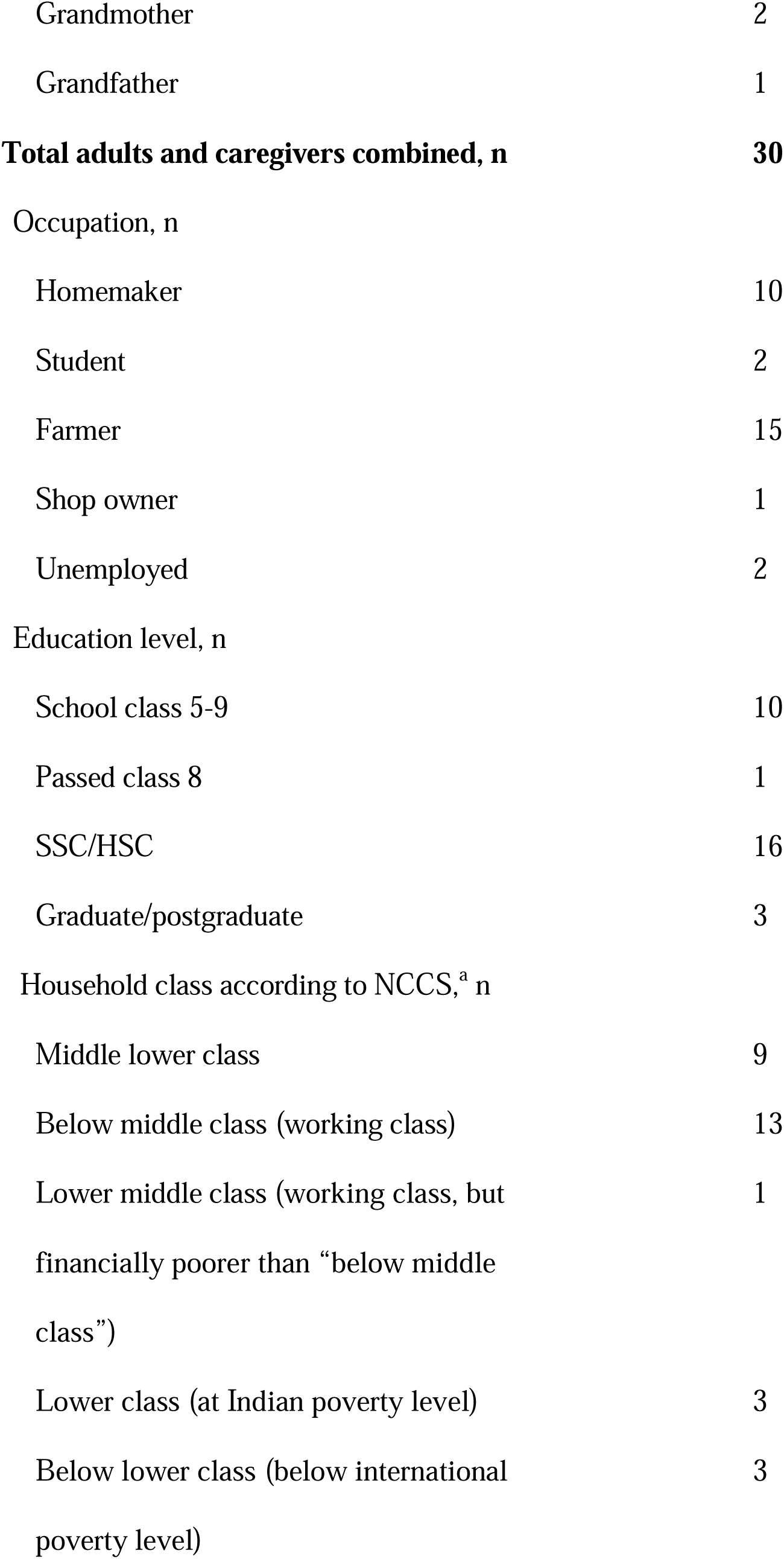
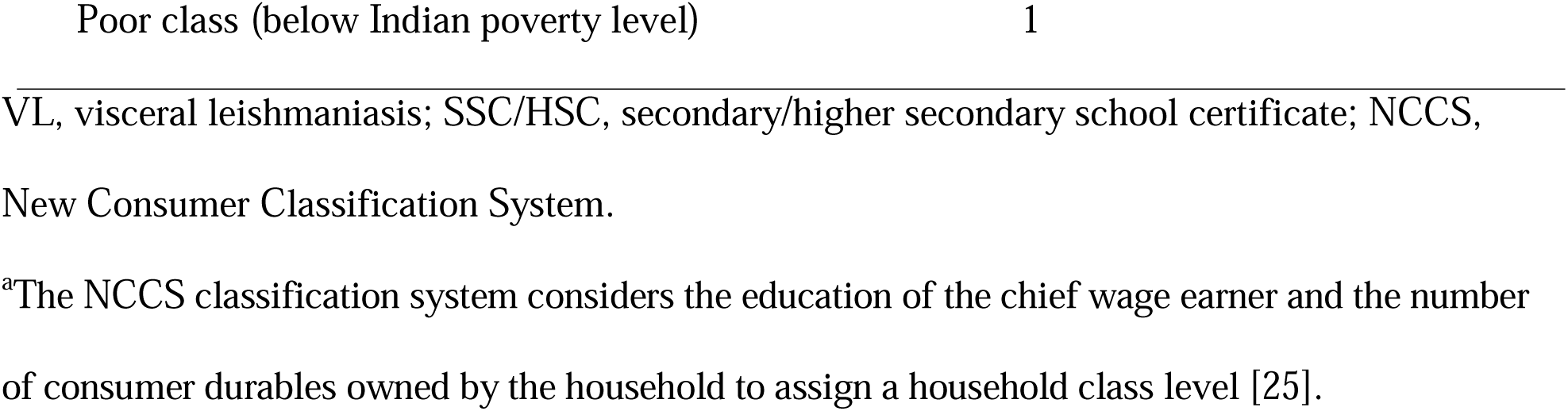
Participant Demographics (N=30)

The overall patient experience could be described as following the path of pre-diagnosis, diagnosis, treatment, and follow-up (**Fig 2**). The timeline for patients ran from Day 0 (onset of symptoms) to first touchpoint (primary care), presentation to diagnosing clinic, receiving diagnosis and treatment, treatment initiated, symptoms resolved, and follow-up (**Fig 3**). Excluding 1 outlier of 90 days, mean time to diagnosis was 13.8 days; range 3 to 21 days; median 7.5 days (**Fig 2**). Including the outlier, mean time to diagnosis was 16.3 days; range 3 to 90 days; median 12 days. No significant differences were observed between males and females (excluding outlier). For adult patients, mean time to diagnosis was 10 days (range 3-21 days; median 8.25 days). Mean time to diagnosis was 19 days (range 5-25 days; median 13.5 days) for pediatric patients (according to caregiver responses).

**Fig 2.**
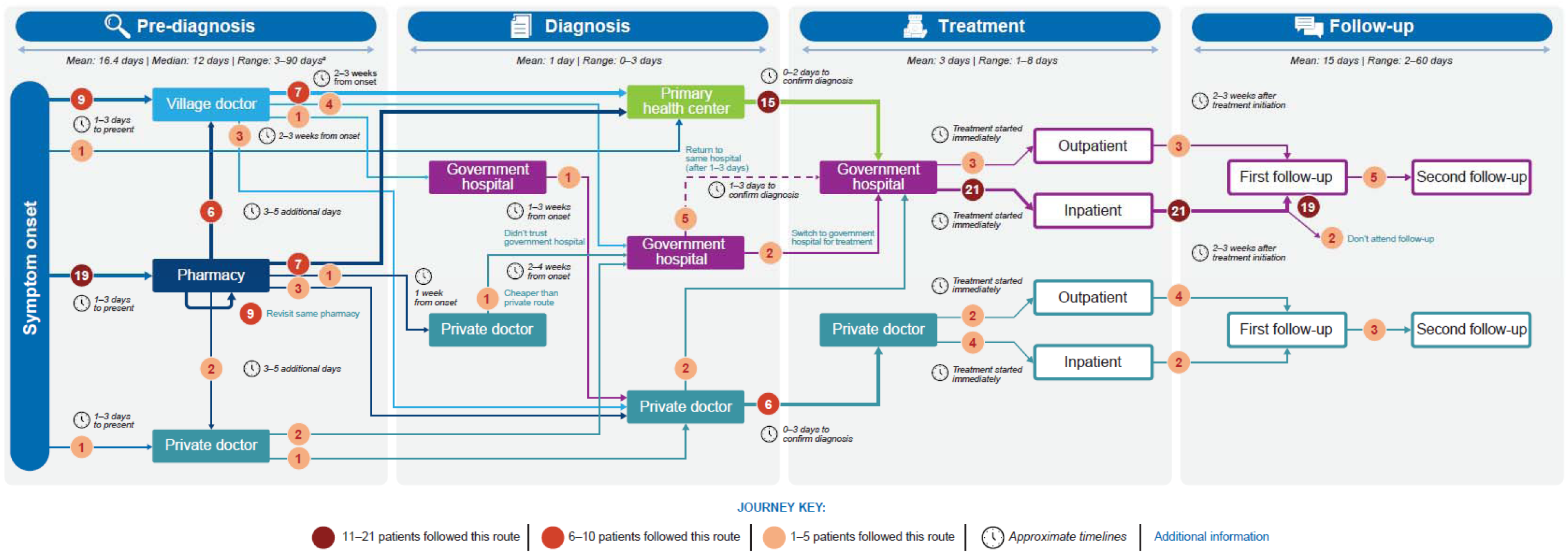
Summary of life experiences of 18 adults and 12 caregivers of children with VL. VL, visceral leishmaniasis. ^a^Excluding 1 anomaly of 90 days. Mean, median, and range breakdowns (with and without outlier) below. - Mean 16.4 days (with outlier); 13.8 days (without outlier). - Median 12 days (with outlier); 7.5 days (without outlier). - Range 3 to 90 days (with outlier); 3 to 21 days (without outlier).

**Fig 3.**
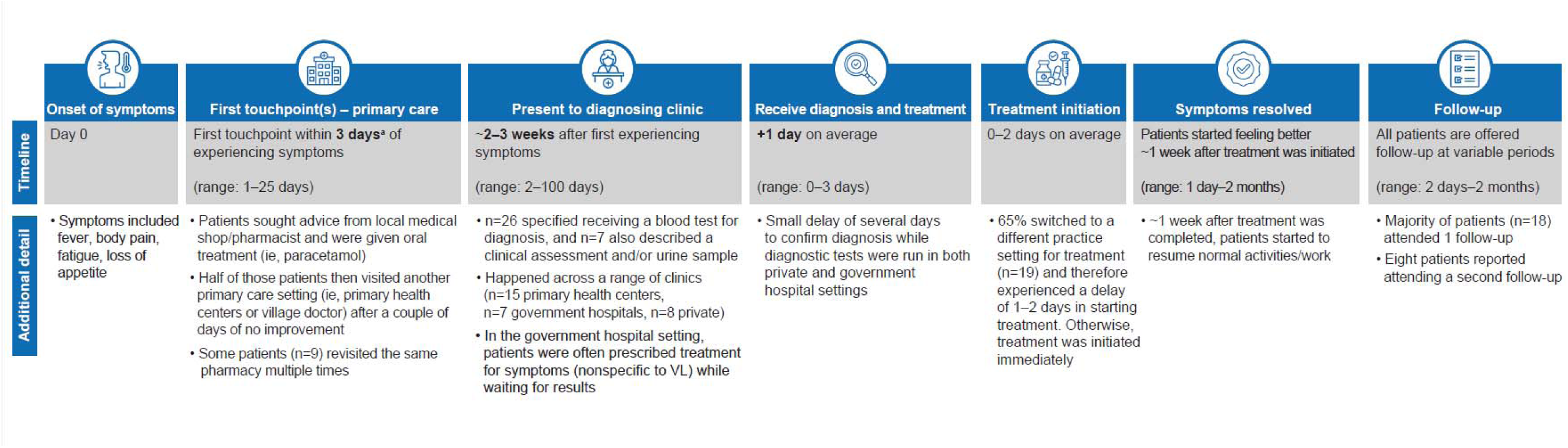
Breakdown of patients’ experiences of VL. VL, visceral leishmaniasis. ^a^Median, 3 days; mean, 3.8 days.

### Patient Life Experience

#### Prediagnosis

VL was suspected at onset of illness by 5 out of 30 participants; of these 5, 2 suspected VL based on suggestion from someone else. The first symptoms experienced associated with VL were often fever and fatigue. As a result of overlapping symptoms with malaria or common seasonal conditions, such as flu, it was common among patients to consider other illnesses before VL. Twenty-one out of 30 patients attempted self-medication and rest upon symptom onset and did not seek treatment until their symptoms persisted for several days. The first point of care was generally a local pharmacist or village doctor (an informal medical practitioner located within a village who fills the gap between local pharmacies and regional hospitals); in these settings, patients often found a low sense of urgency, delaying their diagnosis (**Fig 2**). During this prediagnosis phase, the HCPs, like patients, also considered illnesses other than VL could be responsible. Half of the patients visited a second primary care setting after a couple of days of no improvement (**Fig 3**), and 11 patients remained under the care of nonspecialized HCPs for 2 weeks before VL was diagnosed (**Fig 2**). Delays in seeking diagnosis and treatment also arose from practical barriers for patients and differed between men and women, with women reporting a need for childcare support, whereas men reported the risk of lost wages and a need for time off from work. The challenges patients faced prior to diagnosis included burdensome and debilitating physical symptoms, emotional stressors, absence from work and school, and financial burden.

#### Diagnosis

Upon diagnosis, most patients (n=19; 63%) had to switch to a government hospital for treatment: 15 were referred from a primary health center where treatment for VL was not available, 2 switched from a private doctor for cost reasons, and 2 switched from another government hospital because treatment was not available there (**Fig 2**). These switches caused a 1- to 2-day delay in treatment initiation; thereafter, treatment was started immediately. The time from symptom onset to diagnosis (2-3 weeks in most cases) is in line with the expectations of the Indian National Elimination Program [14]. However, our research shows 7 caregivers of pediatric patients reported taking >2 weeks to present to a diagnostic setting (from 5-9 additional days), due to financial/logistical barriers as well as the desire to seek private care for their child.

#### Treatment

Although treatment is free in a government hospital setting, some patients (n=6; 4 pediatric and 2 adult patients) received treatment in a private healthcare setting (**Fig 2**). All of these patients paid out of pocket at a private hospital due to the fear of long wait times at government hospitals and a lack of trust in the quality of the treatment there. Among the 24 patients who received treatment in a government hospital, 21 were inpatient (treatment duration range, 1-4 days) and 3 were treated as outpatients (treatment duration range, 1-3 days). The majority of patients (n=27) described receiving infusion treatment, but patient descriptions indicated single-dose liposomal amphotericin B may not have been used in some cases. Some patients recalled being treated by injection and/or tablets. Patients began feeling better around 1 week after treatment initiation (range 1 day to 2 months), with most reporting resumption of normal activities approximately 1 week after treatment completion.

The key challenges in this part of the patient life experience were financial burden and the need to travel long distances to obtain treatment. Patients all reported travel burdens; the average reported distance patients needed to travel for treatment was 15 to 20 km (range 3-45 km), with associated financial costs. For those patients who had to return to a distant facility for diagnosis, or those who had to switch treatment facility, the travel burden was even higher. One female adult patient described her experience as “*We delayed the treatment by two weeks because of the financial issue. I faced problem financially because we had to pay the expense of vehicle for travelling. And my school and coaching was getting missed because of my weakness and dizziness.”* Additional challenges for patients and caregivers included unpaid leave from work and finding support for childcare and other responsibilities at home. Furthermore, 8 inpatients reported needing to be accompanied so they could be provided food from outside the hospital during their stay, resulting in costs for food and accommodation near the hospital. As the caregiver of one female pediatric patient noted “*I didn’t spent money in the treatment. But I had to spent money in travelling and etc. Costs getting to hospitals*.”

#### Follow-up

All patients were offered follow-up, ranging between 2 days and 2 months posttreatment. Posttreatment, the majority of patients (n=28) attended the first follow-up appointment and 8 reported attending a second follow-up appointment. For 18 patients, these consultations consisted of a blood test, which they understood was to evaluate for any sign of VL. Thirteen participants or caregivers reported that they were also given medication for what they understood could be to improve their overall health or strength. Follow-up requirements added to the overall financial and time burden experienced by patients with VL, and only 1 patient reported receiving financial reimbursement from the government upon recovery.

### Disease Awareness

Patients and caregivers reported limited awareness and understanding of VL prior to their own experience. Prior to their own or their family member’s diagnosis, the majority of patients and caregivers (>60%) had heard of VL and were familiar with the disease at least by name (kala-azar). Approximately 30% of participants could recognize the seriousness of VL as a disease, but few (<20%) of the patients/caregivers had any understanding of VL in terms of transmission and associated symptoms. One adult female patient noted “*Before, I didn’t know anything, like how it happens and why it happens. I heard [of VL] first time when I suffered from this*,” whilst another adult male patient commented that “*if someone gets too ill then people in the village think that supernatural activity has happened to him and people start to keep distance from that person. That’s why patient doesn’t tell anyone and try to get it cured faster at home*.” After their experience with VL, the majority of patients and caregivers gained a clearer understanding of the disease and the importance of treatment; however, confusion still persisted about disease transmission. They reported feeling motivated to promote awareness of VL and wanting to advise others in their communities to act quickly if they experienced similar symptoms. Patients also were prepared to advise others where to seek treatment to prevent delays.

### Potential New Oral Treatment

Patients expressed a preference for a potential oral treatment over infusions/injections, with anticipation that it could be taken closer to home and reduce out-of-pocket costs. Among adult patients, all respondents would choose oral therapy over that which they received, but some perceived that the tablets would have a slower onset of action and it may take longer to feel better/be cured. As the caregiver of a male pediatric patient stated, “*I would choose 14 days treatment because in this time, the patient would get cured completely and slowly*.” Both treatment durations discussed (7 days and 14 days) were acceptable, with an overall preference for 7 days. Of those who responded, patients stated that they would be willing to pay a maximum of $72 (□5985) from their pocket for their treatment course, with a range of $0.12 per tablet to $72 for the whole course. The high end of this range represents many months of work for patients in lower socioeconomic categories.

Caregivers reported good acceptability of a hypothetical pediatric treatment (capsule containing mini-tablets, which could be added to a spoonful of a child’s food): most think mini-tablets would work for their children. The oral route was preferred versus infusions/injections by 11 out of 12 caregivers to avoid painful injections, and as it would presumably not require HCP administration or travel. Caregivers mentioned they would pay ≤$60 (4987) for this treatment, with a range of $0.50 per tablet to $60 for the whole course. The caregiver who stated a preference for infusion suggested that they felt 1 infusion may be easier to manage than multiple oral treatments.

## DISCUSSION

### Patient Life Experience

In summary, the findings of this study indicate that across the patient life experience, the route to diagnosis of VL was challenging, consisting of multiple touchpoints. Despite the patients’ and caregivers’ rapid presentation to HCPs in their local communities following symptom onset, challenges in making a differential diagnosis, sometimes coupled with a lack of urgency from community HCPs, appeared to contribute to delays in diagnosis and treatment. Further delays resulted when patients were diagnosed in a facility that did not have the appropriate VL treatment available. During the treatment phase, patients faced financial challenges associated with travel to another healthcare facility and were burdened by the need to disrupt work or home life responsibilities during treatment that was often far from home. Patients attended at least one follow-up appointment as suggested. After their experience with VL, patients and caregivers had a better understanding of VL and were motivated to promote awareness of the disease in their local communities.

### Barriers to VL Treatment

Poverty is an overwhelming risk factor for VL and PKDL [26]. Poor housing, malnutrition, and lack of sanitation create and exacerbate barriers to disease care, education, and awareness. Leishmaniasis disease can co-occur with malnutrition and HIV infection, estimated to co-occur in 3.4% of patients with VL in India [27], further complicating diagnosis and treatment. Even when treatment is free, households may experience catastrophic effects of direct and indirect medical costs, compounding existing financial strain in low-income communities. The financial impacts of leishmaniasis are especially burdensome for women, who disproportionately experience poverty [26]. Although the majority of patients (77%) in the current study came from a socioeconomic background between middle lower class and below middle class (working class) categories, they still experienced substantial financial hardship (see **Table 2** for details).

Throughout the VL life experience, the key barriers to treatment identified in this study relate to financial burden, absence from work/home duties, and long-distance travel to hospital settings. The effect of these barriers to treatment was amplified by the need for multiple appointments throughout the patient life experience from prediagnosis through follow-up, often at several different locations. Financial incentives for VL identification, diagnosis, and treatment are offered to HCPs and patients from the National Centre for Vector Borne Diseases Control (NCVBDC) [9, 28].

##### Callout Box 1

Financial Incentives [9, 28] as of December 2023

- ASHAs [29] are offered $6 (lJ500) for case identification, complete treatment and follow-up of each VL or PKDL patient
- One time wage loss incentives/compensation of ($6 [lJ500]) is offered by the NVBDCP to patients with VL [2]
- In addition, the State Governments of Bihar and Jharkhand offer compensation of $79 (lJ6600) and $48 [lJ4000] for each patient with PKDL who completes treatment [2]

Free diagnosis and treatment for VL and PKDL patients are offered at all community health centers and primary healthcare centers in endemic areas. However, although standard-of-care therapy is available for free under the National Elimination Program, with accompanying financial incentives, our research shows there are high out-of-pocket costs for patients along the treatment continuum and a lack of awareness of the incentive program. In the current study, only one patient reported receipt of any reimbursement from the program. The participants’ accounts of financial challenges throughout the treatment process suggest that they encountered financial hardships. Consequently, these results imply that there may be obstacles for patients to benefit from this financial reimbursement program. Also, notably 6 of 30 participants were treated in the private sector, with high associated additional costs (not specified by participants in the current interviews).

### Future Opportunity to Bring Treatment Closer to Patients

Although substantial progress has been made toward elimination of VL in India, challenges remain in traversing the “last mile” toward elimination, including careful surveillance despite rough terrain, low socioeconomic conditions, and lack of awareness [7]. Successful VL elimination and prevention of resurgence will require integration of control efforts into national health programs [10]. A critical action called for in the WHO roadmap is to develop more effective and user-friendly treatment and diagnostics for VL [3] to support the elimination of VL. There is an unmet need for new oral, efficacious, safe, well-tolerated, convenient, short-course, easy-to-use drugs in VL; current research is ongoing to identify and develop new drugs addressing this need. Our research demonstrated that a hypothetical oral treatment for VL would be preferred by 29/30 patients and caregivers due to convenience and at-home administration. Patients considered that an oral treatment would resolve barriers, such as finding childcare while in the hospital, cost of transportation to the hospital, and a partner having to miss work to look after the family. However, it is important to note that the majority of the patients in this study (n=27) came from middle lower to lower middle class backgrounds (working class and above), whereas many patients with VL have traditionally come from more economically disadvantaged backgrounds [26] (See **Table 2**). The unexpectedly high incomes noted here may explain why patients reported being willing to pay up to $72 for a potential course of oral VL treatment.

### VL Elimination in India

In order to achieve the global goal of leishmaniasis elimination by 2030, continued international efforts must address these risk factors and vulnerabilities [3]. Strategies to prevent VL resurgence and support elimination programs are currently a global topic of interest. In 2023, Bangladesh was the first country to eliminate VL as a public health problem [30]. VL elimination in India is considered possible and feasible because VL features focal transmission, does not have an animal reservoir, is transmitted through a single vector, has an effective diagnostic test, and can be effectively treated using a single-dose infusion. Indeed, VL incidence rates in India have been substantially reduced, with the most recent provisional government data reporting only 520 VL cases nationwide in 2023, 329 of which were in Bihar, compared with >9000 cases in 2014 [6].

Our study results suggest a potential target of improvement in the VL patient life experience is reducing time to treatment. Although patients presenting with fever often first seek care from local practitioners in a timely manner, they may experience a substantial delay between symptom onset and diagnosis and subsequent initiation of treatment. Thus, prompt treatment initiation requires awareness and alignment from local private medical practitioners (qualified or unqualified). Apart from the benefit of reducing disease morbidity for the individual, the public health benefit in reducing time to treatment is because VL and PKDL patients serve as potential disease reservoirs in the community. Infection studies have shown that patients with VL or PKDL can pass the infection to sand flies when bitten, continuing the cycle of infection and transmission [31, 32]. VL and PKDL patients were found to be no longer infectious to sand flies by 30 days posttreatment (by microscopy) [32]. An earlier study of the patient’s care-seeking experience in VL in Bangladesh, India, and Nepal argued that improvement in the public health sector is crucial to eliminating VL, with findings strongly adapted to local conditions [33].

### Study Limitations

Study limitations include the challenges of undertaking studies in these areas, such as the low literacy rates in the region; the extremely remote locations with poor transport links that created barriers to healthcare access and also fieldwork; and the potential for recall bias due to time elapsed (≤12 months) since treatment. Because this study was designed to be self-reported, the majority of respondents were not able to recall specific treatment details (frequency and duration); patients were also unable to distinguish VL-specific treatment from supportive treatment. Although many patients described receiving infusion treatment, patient descriptions indicated that the standard-of-care single-dose liposomal amphotericin B may not be used in all cases. Features such as complex treatment regimens, difficulty differentiating supportive versus active therapies, lack of information given to patients about the names of their medications, and concomitant treatments add to the challenges in discerning VL-specific therapy. This confusion prompted the protocol amendment to require documentation of VL diagnosis and treatment after the first 22 completed interviews; however, 18/30 patients could not provide documentation for their VL diagnoses and treatments (they had lost their VL card or were unavailable for follow-up contact). Patients enrolled in this study tended to have a higher income and education level than would be expected among patients with VL in India. Additionally, the caregiver sample reflects only the experience of caregivers of pediatric patients aged 10 to 14 years, although there was an attempt to enroll caregivers of younger pediatric patients.

In contrast to previous studies of VL in India [23, 24, 33, 34] that recruited patients directly from VL hospitals, our study allowed us to identify patients in their local setting and better understand their individual needs and barriers to care. This patient-centric approach is both a limitation and a strength. This is a limitation of the study, as initially we did not have VL cards and confirmation of diagnosis for all patients and needed to add this validation step for previously interviewed patients. However, selecting patients outside the clinic setting revealed a group of patients choosing private health care who would have been missed otherwise. This approach to identifying unmet needs and potential gaps can be useful in furthering the aims of the National Elimination Program for VL.

### Future Research

Building on our research into patient beliefs and experiences in the public and private health sectors when seeking care for VL, future research in this area is needed to understand the segmentation between care in the private and public sectors. Despite an assumption that public health care covers all needs for VL, the extent to which the private sector is utilized in VL care is still not fully understood. Opportunities for collaboration between private and public sectors could therefore be further explored. In addition, considering the previously mentioned lack of uptake in the financial reimbursement program, it may be worthwhile to investigate the obstacles patients encounter in accessing the currently available financial assistance for VL diagnosis and treatment.

Furthermore, as per the diagnostic guidelines in the Indian health care system, VL is suspected in patients following persistent fever (≥14 days) or even earlier when no response has been achieved with antimalarial medications [14]. Our findings indicated that patients sought care much sooner than 14 days after symptom onset, demonstrating an opportunity for earlier diagnosis. Potential adaptations in the current health care system guidelines that encourage earlier testing for VL may result in quicker diagnosis and treatment and reduced patient burden. However, early diagnosis is also limited by available serological diagnostic methods. A validated diagnostic tool with multiple uses for different causes of fever at the point of care such as the development of an antigen-based rapid diagnostic test can be used for both VL diagnosis and determination of cure, would represent a significant advancement in the management of patients with VL.

Finally, leveraging the learnings from this research, it may be fruitful to pursue similar patient experience research in East Africa, a region of high VL incidence and a target of current and future WHO elimination strategies [3].

## CONCLUSIONS

Although in a relatively small sample size, this study reveals barriers to disease awareness and access to care for patients with VL in the Bihar area. Our data demonstrate that even though most patients with VL seek care for symptoms early, diagnosis occurs, on average, 14 days following symptom onset. The current standard-of-care therapy was often unavailable in the health care centers where a patient received their diagnosis, further delaying treatment administration. Oral therapy as an alternative to current parenteral treatment would be preferred both by adult patients and caregivers of pediatric patients and may result in decentralization of treatment and lessening the current financial and travel burden associated with access to the current standard of care.

## Data Availability

Study data are available in Supporting Information 1.

## Acknowledgments

Medical writing support was provided by Katie Gersh, PhD, CMPP, of Lumanity Communications Inc., and was funded by Novartis. The study was conducted by Kadence International, under direction from the authors and funded by Novartis. We gratefully acknowledge contributions of the patients and their families who volunteered to participate in this study.

## Author Contributions

All authors reviewed and approved the manuscript. S. Sundar: lead author, in addition to conceiving, methodology, validation, analysis, and writing. F. Alves: methodology, critical review/editing. K. Ritmeijer: conceptualization, methodology, analysis, writing. M. den Boer: conceptualization and interpretation, critical review/editing. C. Forsyth: methodology, critical review/editing. B. Hughes: design, conceptualization, analysis, and writing. C. Zamble: methodology, execution and analysis of the market research. K. Carter: design, conceptualization, analysis, and writing. G. Angyalosi: conceptualization, design, data interpretation, analysis and writing.

## Notes

### Competing Interest Statement

S. Sundar has received consulting fees from Novartis Pharma AG. F. Alves and C. Forsyth are employed by Drugs for Neglected Diseases initiative (nonprofit organization). K. Ritmeijer and M. den Boer have no conflicts of interest to disclose. B. Hughes is employed by Novartis Pharmaceuticals UK Ltd and received stock options from Novartis Pharma AG. C. Zambles company (Lumanity) has been contracted and paid by Novartis Pharma AG for writing support and market research. K. Carter and G. Angyalosi are employees of Novartis Pharma AG and received stock options from Novartis Pharma AG.

### Funding Statement

Yes

### Author Declarations

Institutional Review Board (IRB) ethics approval was received from Sigma-IRB (https://www.sigma-irb.com/) on December 28, 2022.

