## Supplemental Information 1 for "Patient insights research exploring disease awareness, patient life experience, and current management of visceral leishmaniasis in Bihar, India"

### VL Patient Understanding

#### Data report

September 2023

Lumanity

### Key differences

#### Pre-diagnosis

**Male vs. Female:**

- No differences in timeframe or settings, however, their described experiences differ
- Females experience delay to diagnosis due to child-care support barriers whereas male experiences involve time off work (financial)

**Adult vs. Pediatric:**

- Longer route to diagnosis for pediatric patients (an additional 5 - 9 days vs. adult patients)
  - Mixture of lack of awareness and urgency to escalate (additional 3 days on average to present), and financial/ logistical to travel
  - Additionally, more likely to face delay due to saving for private setting for their child

#### Diagnosis

**Male vs. Female:**

- No difference

**Adult vs. Pediatric:**

- Caregivers are more likely to go to a private doctor for diagnosis (n=4/12) due to wanting quick and ‘trusted’ answers for their child (adults n=2/18)

**Government vs. private:**

- Diagnosis received typically 0-1d from presentation in private setting vs. slightly longer in government setting (1-3d) where patients more likely to receive symptomatic tx whilst waiting for diagnosis
- Cost implications for both – travel for government, consultation/ tests for private

#### Treatment

**Male vs. Females**

- Female patients have additional logistical considerations if an in-patient (e.g., finding beds for chaperones, and child support)

**Adult vs. Pediatric :**

- Pediatric patients more likely to be treated in private setting (n=5/12 peds vs. 2/18 adults)

**Government vs. private**

- Queues associated with government setting to receive treatment

#### Follow up

**Male vs. Female:**

- No difference

**Adult vs. Pediatric:**

- No difference

**Government vs. private:**

- Patients treated in private setting describe greater variability in timeline for follow-up vs. government hospitals (private ranging 8 days to several months; govt. ranging 2 – 20 days)

**Inpatient vs. outpatient treatment setting:**

- Those treated in outpatient setting more likely to describe multiple follow-ups (n=5/7 outpatients vs. n=3/23 inpatients)

Lumanity

### The route to diagnosis is often a long and challenging journey, consisting of multiple touchpoints and failed symptomatic treatments

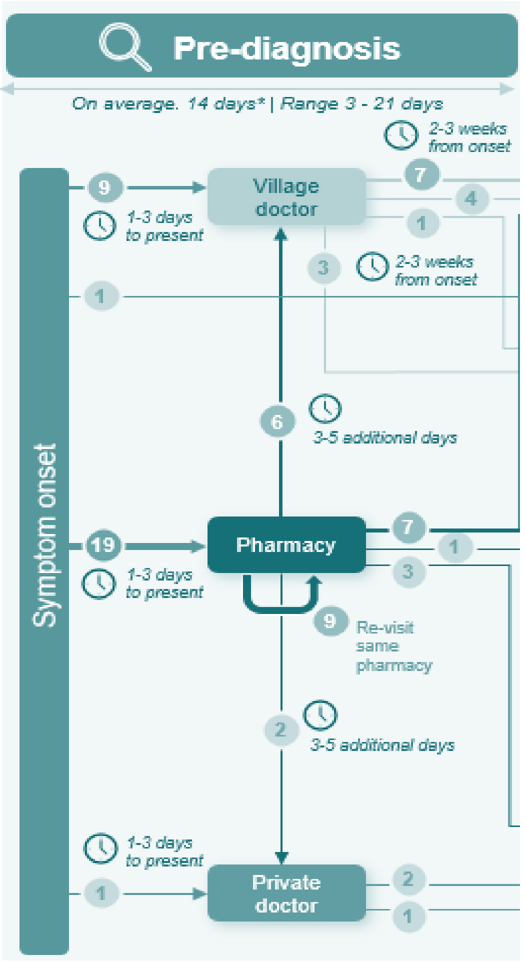


First symptoms experienced are often fever and fatigue, which are at first attributed to conditions like flu

Healthcare advice sought within approximately 3 days after symptom onset, initially from:

- Local pharmacist / medicine shop
- Village doctor or private doctor

Pharmacy / medicine shop, village doctor or private doctor first touchpoint for most patients – where patients receive generic fever treatment, which doesn't offer sustained relief of symptoms

Patients visit 1-2 different settings leading up to diagnosis, sometimes re-visiting the same setting (pharmacy) multiple times

Adult patients (female and male) face similar timelines to diagnosis; but pediatric patients take longer to reach VL diagnosis than adult patients (an additional 5 - 9 days on average)

- Ranging 5d-25d for pediatrics; 3d-21d adult patients

Multiple rounds of failed medications and growing concern and anxiety trigger presentation to a PHC or hospital, where diagnostic tests for VL are done

High socio-economic burden in this phase: patients unable to perform their work / duties, spend money on multiple medications, and experience stress and concern

### Typical first symptoms experienced are fever and fatigue, often misinterpreted as a common or seasonal illness by patients

***Most frequently observed symptoms***

Majority first experience a fever

Accompanied by weakness, fatigue, body pain / cramps, headaches, and lack of appetite, appearing within a couple of days

Fewer mentions of dizziness, stomach-ache, darkening of skin

Minority mentions of brain fog or inability to think clearly, swelling of stomach, irritability

***Least frequently observed symptoms***

***Symptoms range from moderate to severe levels***

Symptoms alone don't immediately drive patients / prompt need for escalation of care to a hospital setting. Despite being burdensome and often debilitating, the condition is not felt to be urgent at this stage

VL is not suspected at this stage based on symptoms alone, due to overlap with a common illness, like flu

Some mentions of patients worrying that they may have coronavirus, malaria, or typhoid

*First of all I experienced fever and I took medicine after 2 or 3 days. Then I experienced headaches and dizziness, then stomachache… I started to fear and got irritable… I thought that it is a normal fever, but I was not getting cured.*

**- Male, Adult patient**

### Patients visit 2-3 healthcare providers seeking medication and advice for their unknown illness, prior to presenting to a diagnostic setting

*Time from symptoms to diagnosis takes ~16 days on average (range: 3d - 3.5months)*

**1**

During first 1-2 days of symptoms, patients self-medicate with paracetamol and rest, and in some cases herbal or homeopathic remedies

When symptoms do not ease, most patients **first visit a pharmacist** (or 'medical shop')

- Typically receive conventional fever medication / further paracetamol
  - Minority specifically recall injections (n=2), antibiotics (n=1)
- Most unable to recall actual treatments given, other than 'fever medicine'

**2**

Symptoms do not improve after 2-3 more days, triggering patients to re-visit their local **pharmacy**, or present to a **new setting in their locality, such as their village doctor**

- Minority visit a private doctor or receive advice from a community healthcare worker at this stage (n=3)
- Patients typically given further fever medication (tablets or syrup) for several days, along with advice to come back / seek further care if not resolved
  - Minority recall being given injections (n=3), antibiotics (n=1)
  - Min. describe tests for other diseases, such as typhoid and malaria

**3**

**For the majority, it is at this stage where need to travel to a PHC / hospital is triggered**

Some patients (n=4) visit a third setting (most commonly a village doctor), continuing to seek care in local area and further delaying presentation to diagnostic setting

### Escalation to diagnostic setting is often delayed due to avoiding travel and added expenses

**A shared patient experience is that a lack of knowledge of VL and its symptoms in the community, including amongst presenting HCPs, causes delay in escalation of care / presentation to place of diagnosis**

#### Additional key barriers include:

**Travel**

- A challenge for many patients, if the hospital is a considerable distance away from their home
  - May require a chaperone (↑female patients); children are always accompanied by caregiver
  - Results in patients cycling through local healthcare settings (i.e., pharmacists / medical shops, village doctors) for longer, delaying presenting to hospital if far away from home

**Childcare**

- Those who have dependent children note this is a reason for delay (↑female patients, caregivers)

**Finances**

- Financial implications of travelling (public transport or car hire)
- If wishing to seek private care for their condition, put it off until they have some funds / until they feel they can't delay any longer (↑ caregivers)

→ Challenges faced more significant for caregivers, with n=7 taking 2+ weeks to present to diagnostic setting due to these barriers (including an additional 5-9 days for first presentation) and due to the desire to seek private care

→ Results in patients persisting with symptoms for longer, despite significant physical impact and worsening quality of life

*[We didn't go to a doctor straight away] due to lack of money for treatment. Then when we did have money for treatment, I took him to a private practice after seeing a government doctor*

**- Caregiver, of Male Pediatric Patient (age range, 11-15 years)**

*I have small children at home, and no one was there at home to take care of the children. It was not easy to go to the hospital with them. My husband had to go to his work. So that's why it took so much time*

**- Female, Adult Patient**

### Patients are reassured at diagnosis regarding curative treatment, but often advised to go elsewhere to receive it

**Doctors convey a sense of reassurance at the point of consultation; they will survive, and they will receive a cure**

**Focus of discussion is treatment options and next steps**

1. **Accessibility**
2. **Inpatient** vs. **outpatient care**
3. Specific nuances for **government** vs. **private** settings

*Minority recall of explaining the cause of VL*

**1 Availability of VL treatment in current setting can prompt a need to switch**

- N=15 in CHC / PHC; patients referred to a government hospital for treatment
- N=1 mention of patient being advised to go to private care (but due to cost impact, went to a government hospital)
- N=1 patient in a govt hospital reported having to switch to another govt hospital due to no treatment available

**2 Inpatient treatment advised (possibility of outpatient treatment)**

- Majority of patients accept being admitted for VL treatment
- N=1 caregiver refuses inpatient care due to cost implication of inpatient stay (in private setting)

**3 Private setting (n=21)**

- Cost of treatment and inpatient stay explained (in addition to diagnosis/ consultation costs)
- Patients are made aware of treatment being free in government setting with n=2 switching for this reasons
- Remaining patients (n=6) continue with private care due to greater trust in quality of care

**Government setting (n=4)**

- Logistics of receiving treatment explained
- Potential queueing for treatment
- Potential risk of access concerns in certain hospitals

**Following diagnosis consultation, n=19** (~63%) **switch to a different setting for treatment**

### Majority receive infusion treatment, as standalone or in combination

**A variety of treatments and combinations of treatments were recalled by respondents for VL specifically**

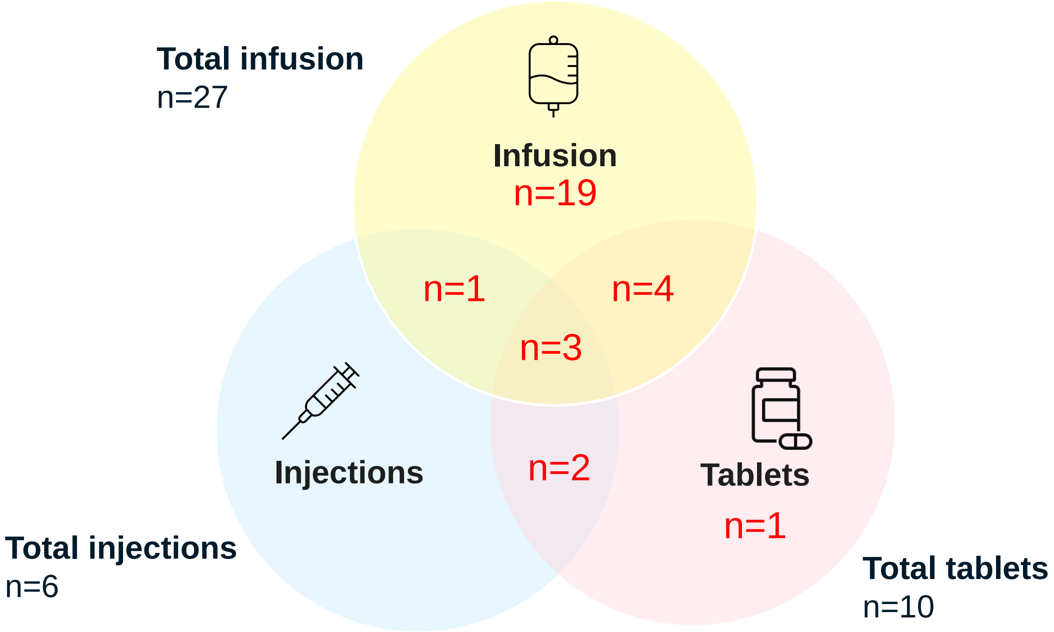


**Other factors also come into play when it comes to what treatment(s) a patient receives**

**VL-specific treatments vs. supportive treatment**

*A mix of infusion, oral medication and/or injection is prescribed for VL specifically, with tonic/syrup or other oral medication mentioned for symptoms such as to regain strength*

**Treatment setting (government vs. private)**

*No difference in treatment observed between government and private setting chooses, minority (n=2) treated in private setting described receiving injections plus tablets / tablets only*

**Inpatient vs outpatient treatment**

*Outpatients more likely to receive injections plus tablets, than inpatients who received infusion*

**Though recollection is not precise, variation in VL treatment regimens is apparent; not all are receiving single dose liposomal amphotericin B**

### Range of duration and types of treatment reported, with gaps in specific details provided by respondents

| Infusion | Injections | Tablets | Tonic/ Syrup |
|---|---|---|---|
| Majority (n=15) patients receive their infusion in the inpatient setting; only n=4 received infusion as outpatients | All patients who received injections with tablets were outpatients | n=1 recalls being prescribed the medication beyond their duration as inpatient | n=6 inpatients and n=5 outpatients also mentioned they received tonic/syrup as a supportive treatment to regain strength |
| Patients recall receiving an infusion for an average of 2.5 hours, but the duration can range from 1-4 hours | Majority (n=4) do not recall the number of injections prescribed single mention of receiving 10 injections | Majority (n=8) patients do not recall how many tablets they were prescribed, minority recall receiving 3 to 4 different types of ('colored') tablets | Few (n=2) patients recall it being prescribed for range of 2 - 4 months |
| Infusions recalled as administered for a range of 1 – 4 days | Few (n=2) recall receiving injections for up to 3 – 5 days, with having to get it twice a day | Wide range in recall of treatment duration: from 5 days to 2-4 months | Single mention of taking supportive treatment for 7 months |

*Additional verbal descriptions were needed from moderators to help respondents differentiate between infusions and injections, and only one patient could spontaneously recall their treatment by name (see details in notes pane).VL treatment cards confirmed L-AMB for n=11 of the respondents, dosage unclear.*

### Patients received similar treatments in government and private setting, however, the degree of financial impact differed

#### Government Setting (n=24)

- Majority of the patients who went to government setting for treatment, chose it due to the treatment being free
- Majority (n=23) of those treated in government setting received infusion
- Majority mention spending on travelling long distance as the government hospitals are far from their villages
- Majority (n=29) make no mention of any government reimbursement schemes, compensation section in n=11 treatment cards left blank
  - n=1 received INR 7000 (85 USD) after recovery
- Most had to wait in long queues to get an appointment with the doctor [single mention of queuing for 2.5 - 3 hrs]

#### Private Setting (n=6)

- All (n=6) chose private setting due to perceived better quality of treatment compared to government setting
  - Majority (n=4) treated in this setting were pediatric patients
- Majority (n=4) of the patients received infusion; n=1 received injections plus tablets; n=1 received only tablets
- These patients experienced heavy financial impact as they had to pay for the treatment, as well as travel
  - n=1 mention of paying around INR 10,000 (120 USD), n=1 mentions paying INR 5000 (61 USD)
  - Single mention of declining treatment in private setting due to cost being approximately INR 200,000 (2400 USD)

*It is good getting free of cost treatment from the government hospital. And if someone will face kala azar in future, then I will suggest to them to go to this hospital and get the free treatment.*

**- Caregiver, of Male Pediatric Patient (age range, 6-10 years)**

*I spent money for travelling and eating food over there. I spent money on extra things.*

**- Male Adult Patient**

*We had taken on debt for getting the treatment done (from private setting).*

**- Caregiver, of Male Pediatric Patient (age range, 11-15 years)**

*Many people said that you don't get right medicine and treatment in government place. I wanted my daughter to get recovered quickly that's why we chose private treatment.*

**- Caregiver, of Female Pediatric Patient (age range, 11-15 years)**

### Patients are satisfied with the treatment experience, but there appear to be gaps on information provided

#### Majority are pleased with treatment offered and readily accept it

Aware their time to diagnosis has been delayed, so relieved to know there is a curative treatment

Respondents respect healthcare professionals' decision on treatment and believe it is the right thing to do

#### Majority recall their treatment experience as good: hospital was hygienic and interactions with medical staff were positive

But majority don't recall being given any information on the purpose of the different treatments

Some of the respondents don't recall being given any information about the treatment risks and/or benefits

Upon treatment initiation, most recall interaction with doctors as involving only dietary advice and sometimes disease prevention guidance

Single mention of nurse trying to speed up infusion process, leaving respondent feeling irritated about treatment experience

*I didn't ask the doctor why are they doing this (infusion as treatment)… No - because doctors know what they are doing.*

**- Caregiver, of Male Paediatric Patient (age range, 11-15 years)**

*No, they did not tell me [at govt hospital] about the expense of the treatment or explain any side effects of the treatment.*

**- Male Adult Patient**

*(Upon treatment initiation) The doctor had asked me to eat simple food, he said not to eat more spicy and oily food for 1.5 months. Then he gave me some medicine. The nurse and doctor told me about the treatment.*

**- Male Adult Patient**

*They told me that I need to get admitted. So they admitted me and gave me an infusion, and then medicine and tonic. They said you will get cure with proper treatment and medicine…They said that infusion and injection are important to be given to me. And they told me about this treatment only.*

***- Male Adult Patient***

### Both inpatients and outpatients faced challenges in receiving the current available treatment

**Inpatients had to be admitted for up to 4 days**

- Some (n=8) had to be accompanied so they can be provided food from outside the hospital
  - n=4 mention their chaperones faced difficulty in finding a place to sleep due to lack of arrangements (↑female patients)
  - Female adult patients with children / female caregivers had to find support for responsibilities at home
  - Male caregivers accompanying their children / male adult patients had to take time off work
- Few mentions of feeling worried about having to miss school, the treatment procedure and loss of appetite because of stress

**Outpatients travelled several times for treatment, for an average of 15-20 Kms**

- n=4 outpatients had to travel an average of 3 times to the hospital to receive rounds of treatment
  - n=1 mentions travelling to and from the treatment location for up to 2 months
  - n=1 mentions travelling twice over a span of 20 days for treatment
- n=1 respondent arranged and paid a local HCP to administer the injections to avoid travelling
- n=1 patient refused to be admitted due to lack of funds

*I used to work in a shop and I had to leave that duty just to take care of my daughter and to take her for the checkup at different places so I faced problems.*
**- Caregiver, Female pediatric patient (age range, 6-10 years)**

*The hospital is too far and I have small children, so I faced problems to reach there. I went with my mother. We had financial problems also, because we had to buy ticket to reach the hospital and come back home. The treatment in the hospital was free but I had to pay the travel costs*

**- Female Adult Patient**

*His father was also stressed and took some holiday from work because we went to the hospital for the treatment.*
**- Caregiver, Male pediatric patient (age range, 11-15 years)**

### During follow-up appointments, patients undergo a blood test and are often advised to continue medication to improve strength

**Most patients recollect starting to feel better within 1 week of receiving treatment**

**Follow-up appointments typically take place at place of treatment, requiring another journey**

All offered at least 1 follow-up, usually after around 15 days after treatment

- Patients examined, usually undergo another blood test
- Patients understanding is that, if the test shows no signs of VL / infection, no further treatment or follow-up is required
- Often prescribed medication (n=13) to improve overall health and strength (new or continuing what was given at VL treatment discharge)
  - Oral medication alone or in combination with a syrup / tonic
- Advised to **seek help if they become ill again** (min. mentions fever specifically)
- Some are also given **lifestyle and prevention advice**, such as managing a healthy diet, and ensuring a clean environment

Some patients (n=8) attend multiple follow-ups (2-3)

- Patients requiring continued symptom management post treatment (e.g., fatigue)
- Mostly those who had longer pre-diagnosis period (2+ weeks), or those who received outpatient treatment

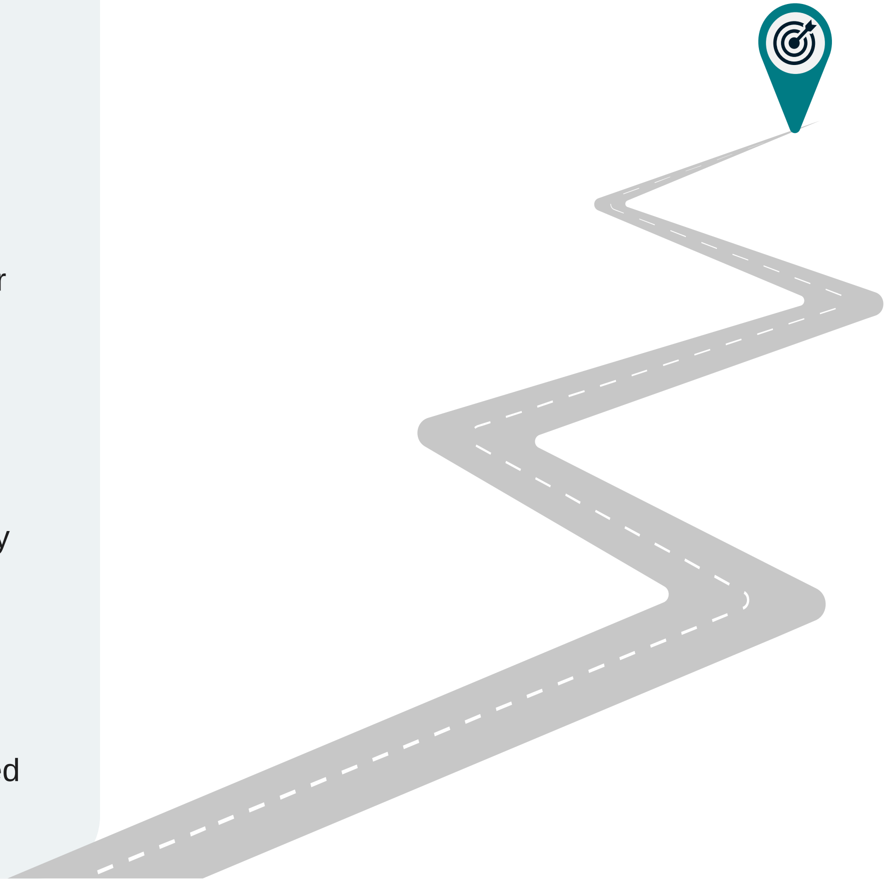

### Requirement to attend follow-up appointments adds to continued financial burden placed on patients, however most patients still ensure they attend

**Financial recovery from patient treatment journey remains burdensome during the physical recovery**

- Continued impact of time off work to attend appointment(s)
- Cost of travel
- Out of pocket expense of continuous treatment and/or follow-up required from private hospitals / clinic*
- Causes significant stress for some, can have serious negative consequences
  - One caregiver deems the inability to pay for private treatment during this period as the reason her child relapsed*
- Need to travel also a burden due to time required, impacting return to normal life
- n=2 caregivers report not taking their child for a follow-up appointment, due to symptoms being resolved and/or due to associated financial burden

Single mention of patient receiving 7000 INR through a government scheme (post treatment at a government hospital)

**→ Despite challenges, almost all patients (n=28) attend at least 1 follow-up appointment, seeming to understand seriousness of condition and need for final sign-off of cure**

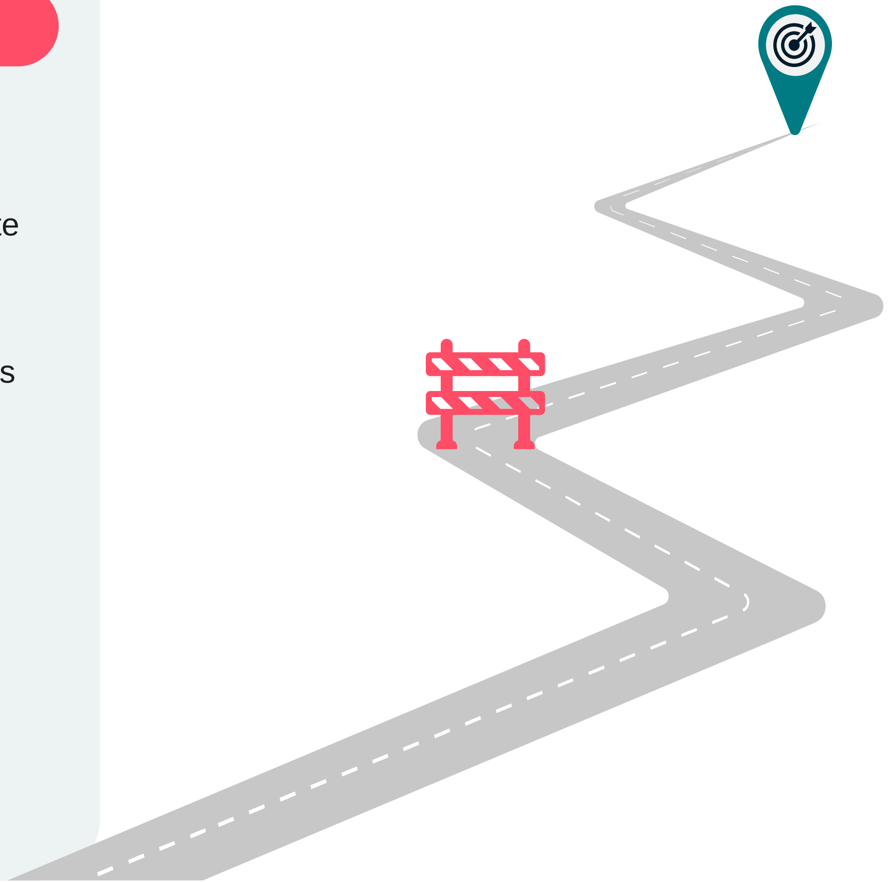



*Additional detail in notes pane

### Participants report that their wellbeing improves after discharge and symptom recovery

**Psychological relief appears to grow as physical wellbeing improves post-treatment**

- **Caregivers: majority report reduced stress** once their child exhibits signs of recovery as they felt it painful to witness their child suffering
- **Patients: sense of support by their family** through treatment & post-discharge - attending appointments and helping with housework
- Treatment is successful in providing a cure: **negative effects** (symptoms, emotional impact) **are relatively short-term; all patients cured of VL**
  - Minority in our sample experience long period of recovery due to ongoing fatigue, impacting quality of life

Majority of patients feel they have **returned to near-normal health by the time of follow-up**, so don't exhibit much anxiety or worry over appointment

- Once VL cure is confirmed at follow-up, it solidifies that life can now go back to "normal" & everything will be "good" now

Patients often **feel motivated to promote awareness of VL** – wanting to advise others in their community to act quickly if similar symptoms experienced, and advising where to seek treatment to prevent delays

***When reflecting on their journey ~1 year after experiencing the condition, sense that negative impacts of VL were temporary for participants – with physical and emotional wellbeing improved, and no significant socio-economic impacts long-term***

### Overall, there is a preference for an oral treatment compared to infusion / injections received

**Hypothetical future treatment discussed:**

- ***A tablet, that would be taken daily, for either 7 days or 14 days, at home following VL diagnosis***
- ***Treatment would be safe, and as effective in curing VL as the treatment previously received***

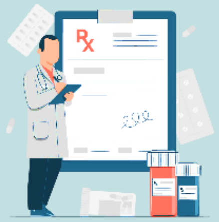

**Positive reactions - preferred formulation**

- If given choice between treatment received and this oral treatment, patients would choose oral (only n=1 caregiver so far would prefer infusion as cured after one administration; don't have to continue with treatments for up to 2 wks which may be difficult for child to maintain)
- Some perceptions tablets would not have as rapid onset of action as injections – will take longer to feel better / be cured
- Respondents would be willing to pay out of pocket for this treatment – between 10 INR per tablet and 6000 IR for the treatment course ($0.12 - $72.17. *Figures not provided by all participants*)

**7 days or 14 days – either option acceptable**

- Overall slight preference for 7 days by majority "If I can get cured in 7 days, why would I choose a 14-day treatment?" - Adult patient. Infers patients would feel better sooner / could resume daily activities sooner.
- Some perceive 14 days as safer; minority concerns that shorter treatment may be more potent, so could have more side effects and would prefer longer duration in that case

**Preferred formulation**

- Oral formulation preferred by majority (n=29) due to convenience and at home administration
  - Respondents mention it will solve challenges such as childcare while in hospital, cost of transportation to hospital and partner having to take time off work to look after the family
- Ultimately the most important thing is being cured of their VL symptoms effectively and safely

### Participant's feedback of hypothetical oral therapy vs. treatment they received

#### Advantages vs. treatment received

- Overall, considered easier and more convenient to most (n=17), because:
  - Able to **administer yourself** vs. injections / IV, which need an HCP
  - **Can take** oral medication **anywhere -** no need to travel / find chaperone, can be done at home
- **No pain -** unlike injections / IV
  - Some report fear (n=5 caregivers, n=6 adult patients) and discomfort due to injections / IV. Children can find injections especially distressing
- **Benefit of simplicity** – requiring only a single medication, so few would prefer due to not having to juggle combination treatments
- **Saves time** vs. injection or infusion and **allows you to continue with life / work** whilst taking the treatment – "*I can't do any work during that [the infusion], and I can't move. But I wouldn't face any problems like that with tablets, and I can go anywhere with the treatment, and I can sit or move according to my will" – Female patient*

#### Drawbacks vs. treatment received

- **Some (n=4)** assume injections provide a 'faster cure' than they perceive an oral medication can offer "*The injection works faster than the tablet, but the tablet is also good"* – Caregiver
- **Single mention** of perceiving oral treatments may be **more likely to cause side-effects**
- Mixed reactions to cost: min.assume this oral medication would cost a lot of money vs. the free treatment they received
  - However, ultimately all (n=30) would pay for a treatment like this, and some assume oral would cost less than their treatment in a private facility or travel to a nearby public hospital

Lumanity

### Caregivers only: Reaction to pediatric formulation

**Hypothetical pediatric formulation discussed:**

- ***A capsule, containing several mini tablets***
- ***Capsule pulled apart, and contents (mini-tablets) added to a spoonful of the child's food***

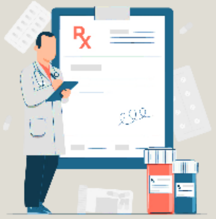

- **Mostly positive reactions**
  - Anticipate good acceptability: most think mini-tablets would work / would be discreet enough for their child
  - Would need to be tasteless: few concerns (minority) child might notice / taste the tablets and not eat food or could cause vomiting
    - Liquid or syrup formulation may be better / more acceptable, which they can mix into milk or honey
    - Or even single capsule easier as don't have to worry about taste (may be due to bias of having older ped sample aged 10-14 years)
- **Oral preferred vs. injections / infusions by most caregivers (11/12)**
  - Keen to avoid painful injections which children don't like,
  - Would not require HCP administration or travel
- **Would pay for a treatment like this**
  - Caregivers mentioned they would pay anywhere between 10 INR per tablet – 5000 INR ($0.50 - $60.15) for this treatment

**Preferred formulation**

- Capsule and mini-tablet formulation is acceptable, and certainly preferred over injections or infusions, but caregivers have mixed opinions over what the ideal formulation would be for their child
  - n=6/12 think syrup / liquid would be best, as easier to administer / higher child acceptability
